# Alcohol use associated alterations in the circulating metabolite profile in the general population and in individuals with major depressive disorder

**DOI:** 10.1101/2023.08.14.23289383

**Authors:** Olli Kärkkäinen, Tommi Tolmunen, Petri Kivimäki, Karoliina Kurkinen, Toni Ali-Sisto, Pekka Mäntyselkä, Minna Valkonen-Korhonen, Heli Koivumaa-Honkanen, Kirsi Honkalampi, Anu Ruusunen, Vidya Velagapudi, Soili M. Lehto

**Author notes:** Correspondence, School of Pharmacy, University of Eastern Finland, P.O. Box 1627, 70211, Kuopio, Finland.

## Abstract

Our aim was to evaluate whether alcohol use is associated with changes in the circulating metabolite profile similar to those present in persons with depression. If so, these findings could partially explain the link between alcohol use and depression. We applied a targeted liquid chromatography mass spectrometry method to evaluate correlates between concentrations of 86 circulating metabolites and self-reported alcohol use in a cohort of the non-depressed general population (GP) (n = 247) and a cohort of individuals with major depressive disorder (MDD) (n = 99). Alcohol use was associated with alterations in circulating concentrations of metabolites in both cohorts. Our main finding was that self-reported alcohol use was negatively correlated with serum concentrations of hippuric acid in the GP cohort. In the GP cohort, consumption of six or more doses per week was associated with low hippuric acid concentrations, similar to those observed in the MDD cohort, but in these individuals it was regardless of their level of alcohol use. Reduced serum concentrations of hippuric acid suggest that already moderate alcohol use is associated with depression-like changes in the serum levels of metabolites associated with gut microbiota and liver function; this may be one possible molecular level link between alcohol use and depression.

## Introduction

Heavy alcohol use and lifetime occurrence of alcohol use disorder have been associated with an increased incidence of a major depressive disorder (MDD) (Grant et al., 2015; McHugh & Weiss, 2019; Odlaug et al., 2016). It has been estimated that around one in six of persons with MDD is suffering from a current alcohol use disorder, and that around every third person with MDD has had a lifetime occurrence of alcohol use disorder (DeVido & Weiss, 2012; Sullivan et al., 2005; Swendsen et al., 1998). However, the relationship between moderate alcohol use and the incidence of MDD remains controversial: some studies show possible small increase in the incidence of MDD (Odlaug et al., 2016), whereas others have not been able to detect any associations (García-Esquinas et al., 2018) or even found evidence for a reduced incidence of MDD (Bellos et al., 2016). Light alcohol use has not been associated with an increased risk of MDD, and light drinkers are commonly used as controls in these studies (Bellos et al., 2016; García-Esquinas et al., 2018; Grant et al., 2015; Odlaug et al., 2016).

One possible mediating effect between alcohol use and MDD are changes in the activity of many metabolic processes. Heavy alcohol use has been associated with an altered metabolome (for a review see Voutilainen and Kärkkäinen, 2019). For example, increased levels of glutamate, citrate, alanine, phenylalanine, tyrosine, glucose, docosahexaenoate, 2- piperidone and phosphatidylcholine diacyls, and decreased levels of glutamine, serotonin, asparagine, hydroxysphingomyelins, gamma-aminobutyric acid (GABA) and phosphatidylcholine acyl-alkyls have been associated with heavy alcohol use (Heikkinen et al., 2019; Jaremek et al., 2013; Kärkkäinen, Farokhnia, et al., 2021; Kärkkäinen, Kokla, et al., 2021; Lehikoinen et al., 2018; Würtz et al., 2016). In a prospective study with an over 30 years’ follow-up, changes in the circulating metabolome, e.g., decreased levels of serotonin and asparagine, preceded the diagnosis of diseases related to alcohol use (Kärkkäinen et al., 2020). Similarly, also MDD has been associated with alterations in circulating levels of metabolites – some of which have also been linked to alcohol use (e.g., glutamate, phenylalanine, citrate and serotonin; for a review see Gadad et al., 2018). This evidence, in addition to the observed overlap in the incidences of MDD and alcohol use, imply that there may well be some shared underlying pathology related to the disturbances in metabolic processes.

We have now investigated whether alcohol use would be associated with similar changes in the circulating metabolite profiles irrespective of the presence of depression by measuring the associations between serum metabolite concentrations and self-reported alcohol use in two cohorts: a non-depressed general population (GP) cohort and a cohort of individuals with MDD.

## Material and methods

### Study cohorts

Two cohorts were used: 1) a general population cohort of non-depressed individuals (GP cohort), and 2) patients with major depressive disorder (MDD cohort) (Ali-Sisto et al., 2016; Kraav et al., 2019). Both cohorts represented the same population, as Kuopio University Hospital from where the MDD cohort was recruited serves the municipality area of Lapinlahti from where the GP cohort was recruited. The Ethics Committee of Hospital District of Northern Savo approved both studies. Participants provided written informed consent before entering the studies.

The non-depressed GP cohort was from a population-based follow-up study consisting of 480 individuals living in the municipality area of Lapinlahti, Finland in 2005. The sample described here was collected as a part of a 5-year follow-up of the study in 2010 (n = 326) but only participants with alcohol use data (n = 289). Further exclusion criteria were a) Beck Depression Inventory (BDI) score of ≥ 10 at the time of the study or 5 years earlier (Beck, 1961), or b) use of medication for depression. After exclusions, a total of 247 subjects were included in the study (Table 1).

**Table 1:**
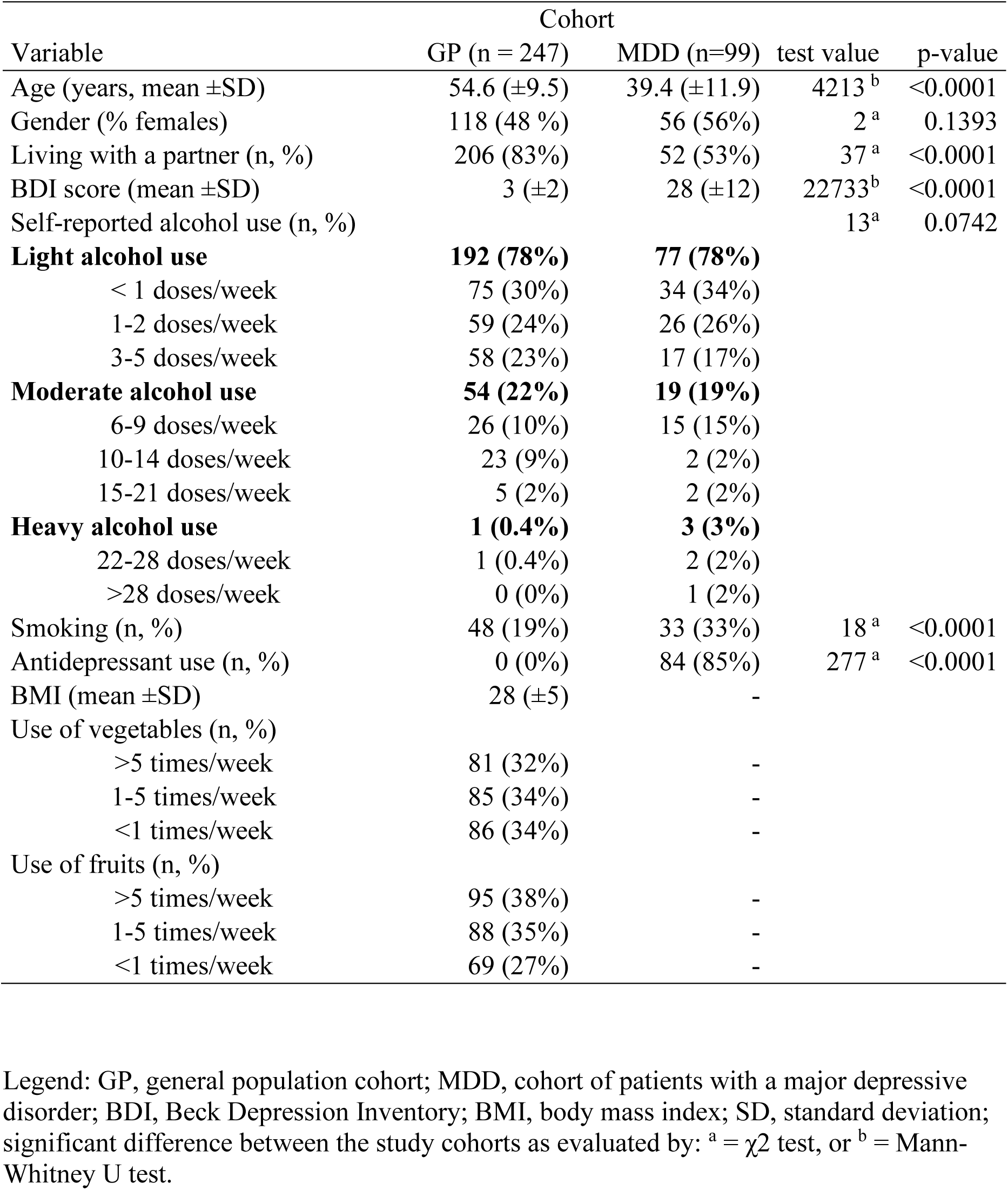
Background characteristics of the study cohorts.

The MDD cohort consisted of 99 outpatients with MDD who were recruited from the Department of Psychiatry of Kuopio University Hospital, Finland. The diagnosis of MDD was confirmed with the structured clinical interview for DSM-IV (SCID) (DSM-IV; American Psychiatric Association, 1994). The exclusion criteria were a) bipolar disorder, b) psychotic disorders, c) a symptomology of mental illness due to alcohol or substance abuse-related diagnosis, d) epilepsy, and e) current somatic conditions preventing participation in the study visits. Most of the participants in the MDD cohort were being administered some form of psychotropic medication: 81 had been prescribed with an antidepressant medication and 48 with an antipsychotic drug with some indication other than a psychotic disease (the medication use has been described in detail in Ali-Sisto, et al. 2016).

### Alcohol use

The participants in both cohorts completed self-report questionnaires from which the weekly alcohol use was extracted. Response alternatives were: < 1 drink/week, 1-2 drink(s)/week, 3- 5 drinks/week, 6-9 drinks/week, 10-14 drinks/week, 15-21 drinks/week, 22-28 drinks/week, or > 28 drinks/week. One standard drink in Finland is defined as a drink containing 12 grams of ethanol. Values higher than 21 drinks per week were considered heavy use, between 6 and 21 drinks per week moderate use, and below 6 drinks per week light drinking.

### Background data, depressive symptoms and psychiatric medications

Background data was collected from the questionnaires completed by the participants. Depressive symptoms were assessed with the Beck Depression Inventory (Beck, 1961) in both cohorts. The use of prescription and over-the-counter medications was also recorded with a questionnaire and confirmed from the prescription documents that the participants provided. The following variables were also collected from both cohorts: age (in years), gender (female/male), regular smoking (yes/no), cohabitation status (married or living with a partner vs. living alone), and education level (higher education yes/no). Use of vegetables and use of fruits (< 1 time/week, 1-5 times/week, > 5 times/week, according to Savolainen et al. 2014), and body mass index (BMI) were only available in the GP cohort.

### Metabolite profiling analysis

Venous blood samples were obtained via venipuncture by trained laboratory personnel and centrifuged to separate the serum. Before venipuncture, the participants were instructed to fast for 12 hours. Serum samples were stored frozen at -80 °C until analyzed. The samples were analyzed in the metabolomics unit of the Institute of Molecular Medicine Finland (FIMM) in Helsinki, Finland. Metabolite profiling analysis of the samples was performed using liquid chromatography-mass spectrometry. Briefly, 10 µL of labeled internal standard mixture was added to the 100 µL of samples, and the samples were allowed to equilibrate with the internal standards. A total of 400 µL of extraction solvent (1% formic acid in acetonitrile) was added and the collected supernatant was dispensed into an OstroTM 96-well plate (Waters Corporation, Milford, USA) and then filtered by applying a vacuum at a delta pressure of 300–400 mbar for 2.5 minutes on a Hamilton robot’s vacuum station. After this, 5 μL of filtered sample extract was injected into an Acquity UPLC system coupled to a Xevo® TQ-S triple quadrupole mass spectrometer (Waters Corporation, Milford, MA, USA), which was operated in both positive and negative polarities with a polarity switching time of 20 msec for metabolite separation and quantification. The Multiple Reaction Monitoring (MRM) acquisition mode was selected for the quantification of the metabolites. MassLynx 4.1 software was used for data acquisition, data handling, and instrument control. The data were processed using TargetLynx software.

### Statistics

In the comparison of the background characteristics of the cohorts, we used Mann-Whitney U test for continuous and χ^2^ test for binomial variables.

For the metabolite profiling data, only metabolites that were above the quantification limits in at least 50% of subjects in at least one cohort were included in the study. Metabolite concentrations below the quantification limit were excluded from the analysis, with the exception of cotinine, where concentrations below the quantification limit were considered to be zero.

In the statistical analysis of the metabolite data, the self-reported weekly alcohol drinking levels were coded numerically (1 = < 1 drink/week, 2 = 1–2 drink(s)/week, 3 = 3-5 drinks/week, 4 = 6–9 drinks/week, 5 = 10–14 drinks/week, 6 = 15–21 drinks/week, 7 = 22–28 drinks/week, and 8 = > 28 drinks/week) to allow us to conduct a regression analysis. We analyzed the two cohorts separately for changes in the metabolite concentrations associated with alcohol use. This was done to focus the analysis on alcohol use related changes, not to changes associated with MDD, which have been reported earlier (Ali-Sisto et al. 2016). We used linear regression to investigate correlations between self-reported weekly alcohol drinking and serum metabolite concentrations. To account for multiple testing, we undertook a principal component analysis and used the number of principal components needed to explain 95% of variance in the datasets to adjust the α-level using Bonferroni’s method to 0.001 (46 and 41 principal components were needed to explain 95% of variance in the GP and MDD cohorts, respectively). Metabolites with p-values between 0.05 and 0.001 were considered as trends.

For metabolites with a p-value below 0.05 in the initial analysis, we conducted a post-hoc analysis using linear regression with possible confounders previously associated with alcohol use and/or metabolite levels (age, sex, use of vegetables, use of fruits, smoking, BMI, education level, and cohabitation status) as covariates. We used JASP (version 0.14), SIMCA (version 17, Umetrics), and Prism (GraphPad Software Inc., version 7) for statistical analyses, multivariate analysis, and figure preparation, respectively.

## Results

The characteristics of the study participants in both cohorts are presented in Table 1. The GP cohort was older than the MDD cohort. With respect to the selection criteria, as would be expected, the MDD cohort had higher BDI scores when compared to the non-depressed GP cohort. Subjects in the MDD cohort were living alone and smoked regularly more often when compared to the GP cohort.

The correlations found between self-reported alcohol use and serum metabolite concentrations with p-value below 0.05 in at least one cohort are shown in Figure 1. However, at the multiple testing adjusted α level of 0.001, the only significant correlations were observed between self-reported alcohol use and serum concentrations of hippuric acid and chenodeoxycholic acid in the GP cohort, and xanthosine in the MDD cohort (Figure 1). Correlations with p-values between 0.05 and 0.001 should be considered as trends that need to be validated in larger cohorts. The correlations between self-reported alcohol use and all 86 measured metabolites are shown in the Supplementary Table 1.

**Figure 1:**
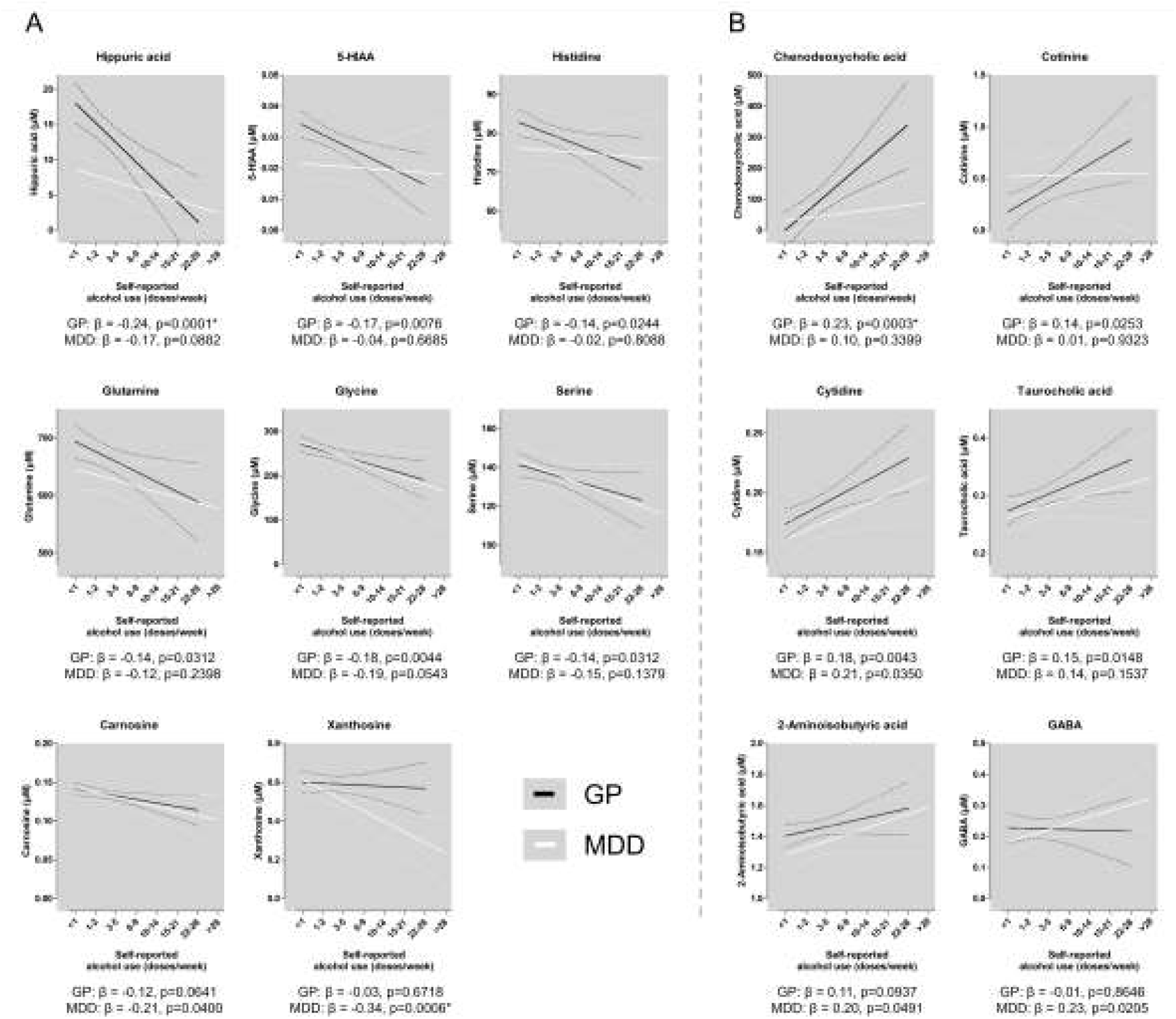
Metabolites associated with self-reported alcohol use. Standardized β coefficients are shown (with 95% confidence intervals as dashed lines) for linear regression models between measured metabolites and self-reported alcohol use with negative (A) or positive (B) correlations. After correction for multiple testing, only correlations between self-reported alcohol use and chenodeoxycholic acid, hippuric acid and xanthosine can be viewed as significant (p-values below multiple testing adjusted α of 0.001). Other correlations should be considered as trends. Legend: GP, general population cohort; MDD, major depressive disorder cohort; β, standardized β coefficient from linear regression model; p, p-value from linear regression model; *, p-value < 0.001 (Bonferroni adjusted α level).

A post-hoc analysis was conducted using linear regression with potential confounders. In the GP cohort, most of the correlations between self-reported alcohol use and metabolite levels remained significant after taking into account possible confounders (hippuric acid β = -0.19, p = 0.0037; chenodeoxycholic acid β = 0.19, p = 0.0049; cytidine β = 0.22, p = 0.0011; 5- HIAA β = -0.16, p = 0.0183; taurocholic acid β = 0.22, p = 0.0012; glutamine β = -0.20, p = 0.0040; and serine β = -0.16, p = 0.0218). There were two exceptions; glycine (β = -0.12, p = 0.0785) and cotinine (β = 0.03, p = 0.5005). Instead of alcohol use, sex correlated more with the glycine concentration (β = -0.15 and p = 0.0382) and smoking status more with cotinine concentrations (β = 0.74, p < 0.0001). In the MDD cohort, all metabolites with a p-value below 0.05 in the initial analysis also had a p-value below 0.05 in the linear regression models when lifestyle factors were included as covariates (xanthosine β = -0.38, p = 0.0002; GABA β = 0.25, p = 0.0186; cytidine β = 0.21, p = 0.0388; carnosine β = -0.21, p = 0.0348; and 2-aminoisobutyric acid β = 0.22, p = 0.0336).

## Discussion

Our main finding was that in the GP cohort, at least moderately increased alcohol use was associated with lower serum concentrations of hippuric acid. The hippuric acid concentrations in those individuals who reported six or more alcohol doses per week were similar to the concentrations observed in the MDD cohort regardless of their level of alcohol use (Figure 1). These results are in line with previous reports, where decreased levels of hippuric acid have been associated with alcohol use, depression, anxiety disorders, and stress (J. Chen et al., 2018; Kärkkäinen et al., 2020; Phua et al., 2015; P. Zheng, Wang, et al., 2013; P. Zheng, Chen, et al., 2013). Although rather little is known to explain how hippuric acid might influence behavior, previous studies have found that low hippuric acid levels may impair neuronal development in fetal mouse brain (Ahmed et al., 2022; Vuong et al., 2020). Moreover, in humans, low serum hippuric acid concentrations have been associated with mild cognitive impairment, whereas high levels of hippuric acid have been linked with beneficial effects on cognitive performance (Rutledge et al., 2021; Yilmaz et al., 2020).

In the GP cohort, similar trends were also seen in the association between self-reported alcohol use and concentrations of 5-HIAA and histidine. Interestingly, the composition of gut microbiota may influence the serum levels of the following three compounds, i.e., hippuric acid (Behr et al., 2017; Fujisaka et al., 2018; Lees et al., 2013; Pessa-Morikawa et al., 2022; Vuong et al., 2020; Williams et al., 2010), 5-HIAA (Lukić et al., 2019), and histidine (H. Chen et al., 2019; Kawase et al., 2017). Moreover, both alcohol use and MDD have been associated with an altered composition and changes in the function of the gut microbiota (Kelly et al., 2016; Leclercq et al., 2014; Llopis et al., 2016; Winter et al., 2018; P. Zheng et al., 2016). Therefore, alterations in the gut microbiota could explain the correlations between serum levels of these metabolites, alcohol use and depression.

However, in the case of hippuric acid, the gut microbiota first makes benzoic acid from phenolic compounds in the diet, and then the liver forms hippuric acid by combining benzoic acid with glycine (Lees et al., 2013). The declines in serum hippuric acid concentrations could therefore also be a reflection of disturbed liver function since decreased hippuric acid production has been associated with alcohol use and impaired liver function (van Sumere et al., 1969). Furthermore, the intake of vegetables or fruits, the main sources of phenolic compounds in the diet, did not significantly correlate with serum hippuric acid concentrations in the present study. Therefore, differences in the intakes of food items with phenolic compounds are not a likely explanation for the changes we observed in serum levels of hippuric acid. More research is needed to clarify the role of hippuric acid in alcohol use and depression.

Moreover, we also observed a positive correlation between self-reported alcohol consumption and serum concentrations of two bile acids, chenodeoxycholic acid and taurocholic acid. These correlations could be associated with altered bile acid synthesis or metabolism, which can also be linked with the gut microbiota (Bajaj, 2019). Increases in serum levels of bile acids have been associated with the organ damage induced by alcohol consumption (Bajaj, 2019; Milstein et al., 1976; Trinchet et al., 1994). Interestingly, there was no correlation between self-reported alcohol use and chenodeoxycholic acid levels in the MDD cohort, which could be associated with altered cholesterol metabolism in MDD since cholesterol is a starting precursor for bile acid formation (Kraav et al., 2019; Lehto et al., 2010). Moreover, in the MDD cohort, we observed a negative correlation between xanthosine and self-reported alcohol use, which was not observed in the GP cohort. This difference is likely linked to the previously reported dysregulation of purine metabolism in the same cohort (Ali-Sisto et al., 2016).

Furthermore, there were several metabolites, which were positively (cytidine, taurocholic acid) or negatively (glycine, serine and glutamine) correlated with self-reported alcohol use in both the GP and MDD cohorts with similar standardized β coefficients (Figure 1). However, due to the smaller sample size, these correlations were not statistically significant in the MDD cohort. These alterations are in line with results from previous studies investigating changes associated with alcohol use in the human circulating metabolome (Kärkkäinen et al., 2020; Lehikoinen et al., 2018; Voutilainen & Kärkkäinen, 2019; Würtz et al., 2016).

Limitations of the study include the relatively small number of subjects in the MDD cohort. Based on the present results, moderate alcohol use seems to be associated with small-to-medium effect size changes in the serum metabolite profile and therefore future studies should examine larger cohorts. We estimated that a sample size of 315 would be needed to observe small-to-medium effect size (f^2^ > 0.1) correlations between alcohol use and metabolite concentrations with the corrected α level used in the present study (0.0001) with 0.95 statistical power. Furthermore, future studies should also use other biomarkers such as phosphatidylethanols, to assess recent alcohol use instead of relying solely on self-reporting (Y. Zheng et al., 2011).

The present results show that already moderate alcohol use is associated with depression-like changes in the serum levels of a metabolite, hippuric acid, which has been linked with both the gut microbiota and liver function. This indicates that changes in the metabolites associated with the gut microbiota and liver function could be one of the molecular level links between alcohol use and depression, although further research will be needed to verify this hypothesis.

## Acknowledgements

The authors wish to thank Dr. Jorma Savolainen for his significant contribution in the collection of the Lapinlahti Study sample, and Dr. Ewen MacDonald for proofreading.

## Data availability statement

The data that support the findings of this study are available on request from the corresponding author, OK. The data are not publicly available due to their containing information that could compromise the privacy of research participants. The study plan approved by the ethical committee and the participant consent terms preclude public sharing of these sensitive data, even in anonymized form.

## References

Ahmed, H., Leyrolle, Q., Koistinen, V., Kärkkäinen, O., Layé, S., Delzenne, N., & Hanhineva, K. (2022). Microbiota-derived metabolites as drivers of gut-brain communication. Gut Microbes, 14(1), 2102878. https://doi.org/10.1080/19490976.2022.2102878

Ali-Sisto, T., Tolmunen, T., Toffol, E., Viinamäki, H., Mäntyselkä, P., Valkonen-Korhonen, M., Honkalampi, K., Ruusunen, A., Velagapudi, V., & Lehto, S. M. (2016). Purine metabolism is dysregulated in patients with major depressive disorder. Psychoneuroendocrinology, 70, 25–32. https://doi.org/10.1016/j.psyneuen.2016.04.017

Bajaj, J. S. (2019). Alcohol, liver disease and the gut microbiota. Nature Reviews. Gastroenterology & Hepatology, 16(4), 235–246. https://doi.org/10.1038/s41575-018-0099-1

Beck, A. T. (1961). An Inventory for Measuring Depression. Archives of General Psychiatry, 4(6), 561. https://doi.org/10.1001/archpsyc.1961.01710120031004

Behr, C., Kamp, H., Fabian, E., Krennrich, G., Mellert, W., Peter, E., Strauss, V., Walk, T., Rietjens, I. M. C. M., & van Ravenzwaay, B. (2017). Gut microbiome-related metabolic changes in plasma of antibiotic-treated rats. Archives of Toxicology, 91(10), 3439–3454. https://doi.org/10.1007/s00204-017-1949-2

Bellos, S., Skapinakis, P., Rai, D., Zitko, P., Araya, R., Lewis, G., Lionis, C., & Mavreas, V. (2016). Longitudinal association between different levels of alcohol consumption and a new onset of depression and generalized anxiety disorder: Results from an international study in primary care. Psychiatry Research, 243, 30–34. https://doi.org/10.1016/j.psychres.2016.05.049

Chen, H., Nwe, P.-K., Yang, Y., Rosen, C. E., Bielecka, A. A., Kuchroo, M., Cline, G. W., Kruse, A. C., Ring, A. M., Crawford, J. M., & Palm, N. W. (2019). A Forward Chemical Genetic Screen Reveals Gut Microbiota Metabolites That Modulate Host Physiology. Cell, 177(5), 1217–1231.e18. https://doi.org/10.1016/j.cell.2019.03.036

Chen, J., Bai, S.-J., Li, W., Zhou, C., Zheng, P., Fang, L., Wang, H., Liu, Y., & Xie, P. (2018). Urinary biomarker panel for diagnosing patients with depression and anxiety disorders. Translational Psychiatry, 8(1), 192. https://doi.org/10.1038/s41398-018-0245-0

DeVido, J. J., & Weiss, R. D. (2012). Treatment of the depressed alcoholic patient. Current Psychiatry Reports, 14(6), 610–618. https://doi.org/10.1007/s11920-012-0314-7

Fujisaka, S., Avila-Pacheco, J., Soto, M., Kostic, A., Dreyfuss, J. M., Pan, H., Ussar, S., Altindis, E., Li, N., Bry, L., Clish, C. B., & Kahn, C. R. (2018). Diet, Genetics, and the Gut Microbiome Drive Dynamic Changes in Plasma Metabolites. Cell Reports, 22(11), 3072–3086. https://doi.org/10.1016/j.celrep.2018.02.060

Gadad, B. S., Jha, M. K., Czysz, A., Furman, J. L., Mayes, T. L., Emslie, M. P., & Trivedi, M. H. (2018). Peripheral biomarkers of major depression and antidepressant treatment response: Current knowledge and future outlooks. Journal of Affective Disorders, 233, 3–14. https://doi.org/10.1016/j.jad.2017.07.001

García-Esquinas, E., Ortolá, R., Galán, I., Soler-Vila, H., Laclaustra, M., & Rodríguez-Artalejo, F. (2018). Moderate alcohol drinking is not associated with risk of depression in older adults. Scientific Reports, 8(1), 11512. https://doi.org/10.1038/s41598-018-29985-4

Grant, B. F., Goldstein, R. B., Saha, T. D., Chou, S. P., Jung, J., Zhang, H., Pickering, R. P., Ruan, W. J., Smith, S. M., Huang, B., & Hasin, D. S. (2015). Epidemiology of DSM-5 Alcohol Use Disorder: Results From the National Epidemiologic Survey on Alcohol and Related Conditions III. JAMA Psychiatry, 72(8), 757. https://doi.org/10.1001/jamapsychiatry.2015.0584

Heikkinen, N., Kärkkäinen, O., Laukkanen, E., Kekkonen, V., Kaarre, O., Kivimäki, P., Könönen, M., Velagapudi, V., Nandania, J., Lehto, S. M., Niskanen, E., Vanninen, R., & Tolmunen, T. (2019). Changes in the serum metabolite profile correlate with decreased brain gray matter volume in moderate-to-heavy drinking young adults. Alcohol, 75, 89–97. https://doi.org/10.1016/j.alcohol.2018.05.010

Jaremek, M., Yu, Z., Mangino, M., Mittelstrass, K., Prehn, C., Singmann, P., Xu, T., Dahmen, N., Weinberger, K. M., Suhre, K., Peters, A., Döring, A., Hauner, H., Adamski, J., Illig, T., Spector, T. D., & Wang-Sattler, R. (2013). Alcohol-induced metabolomic differences in humans. Translational Psychiatry, 3(7), e276–e276. https://doi.org/10.1038/tp.2013.55

Kärkkäinen, O., Farokhnia, M., Klåvus, A., Auriola, S., Lehtonen, M., Deschaine, S. L., Piacentino, D., Abshire, K. M., Jackson, S. N., & Leggio, L. (2021). Effect of intravenous ghrelin administration, combined with alcohol, on circulating metabolome in heavy drinking individuals with alcohol use disorder. *Alcoholism*, Clinical and Experimental Research, 45(11), Article 11. https://doi.org/10.1111/acer.14719

Kärkkäinen, O., Klåvus, A., Voutilainen, A., Virtanen, J., Lehtonen, M., Auriola, S., Kauhanen, J., & Rysä, J. (2020). Changes in circulating metabolome precede alcohol-related diseases in middle-aged men: A prospective population-based study with a 30-year follow-up. Alcoholism: Clinical and Experimental Research, 44, 2457–2467. https://doi.org/10.1111/acer.14485

Kärkkäinen, O., Kokla, M., Lehtonen, M., Auriola, S., Martiskainen, M., Tiihonen, J., Karhunen, P. J., Hanhineva, K., & Kok, E. (2021). Changes in the metabolic profile of human male postmortem frontal cortex and cerebrospinal fluid samples associated with heavy alcohol use. *Addiction Biology*, e13035. https://doi.org/10.1111/adb.13035

Kawase, T., Nagasawa, M., Ikeda, H., Yasuo, S., Koga, Y., & Furuse, M. (2017). Gut microbiota of mice putatively modifies amino acid metabolism in the host brain. British Journal of Nutrition, 117(6), 775–783. https://doi.org/10.1017/S0007114517000678

Kelly, J. R., Borre, Y., O’ Brien, C., Patterson, E., El Aidy, S., Deane, J., Kennedy, P. J., Beers, S., Scott, K., Moloney, G., Hoban, A. E., Scott, L., Fitzgerald, P., Ross, P., Stanton, C., Clarke, G., Cryan, J. F., & Dinan, T. G. (2016). Transferring the blues: Depression-associated gut microbiota induces neurobehavioural changes in the rat. Journal of Psychiatric Research, 82, 109–118. https://doi.org/10.1016/j.jpsychires.2016.07.019

Kraav, S.-L., Tolmunen, T., Kärkkäinen, O., Ruusunen, A., Viinamäki, H., Mäntyselkä, P., Koivumaa-Honkanen, H., Valkonen-Korhonen, M., Honkalampi, K., Herzig, K.-H., & Lehto, S. M. (2019). Decreased serum total cholesterol is associated with a history of childhood physical violence in depressed outpatients. Psychiatry Research, 272, 326–333. https://doi.org/10.1016/j.psychres.2018.12.108

Leclercq, S., Matamoros, S., Cani, P. D., Neyrinck, A. M., Jamar, F., Stärkel, P., Windey, K., Tremaroli, V., Bäckhed, F., Verbeke, K., de Timary, P., & Delzenne, N. M. (2014). Intestinal permeability, gut-bacterial dysbiosis, and behavioral markers of alcohol-dependence severity. Proceedings of the National Academy of Sciences, 111(42), E4485–E4493. https://doi.org/10.1073/pnas.1415174111

Lees, H. J., Swann, J. R., Wilson, I. D., Nicholson, J. K., & Holmes, E. (2013). Hippurate: The Natural History of a Mammalian–Microbial Cometabolite. Journal of Proteome Research, 12(4), 1527–1546. https://doi.org/10.1021/pr300900b

Lehikoinen, A. I., Kärkkäinen, O. K., Lehtonen, M. A. S., Auriola, S. O. K., Hanhineva, K. J., & Heinonen, S. T. (2018). Alcohol and substance use are associated with altered metabolome in the first trimester serum samples of pregnant mothers. European Journal of Obstetrics & Gynecology and Reproductive Biology, 223, 79–84. https://doi.org/10.1016/j.ejogrb.2018.02.004

Lehto, S. M., Niskanen, L., Tolmunen, T., Hintikka, J., Viinamäki, H., Heiskanen, T., Honkalampi, K., Kokkonen, M., & Koivumaa-Honkanen, H. (2010). Low serum HDL-cholesterol levels are associated with long symptom duration in patients with major depressive disorder: HDL-C and the duration of depression. Psychiatry and Clinical Neurosciences, 64(3), 279–283. https://doi.org/10.1111/j.1440-1819.2010.02079.x

Llopis, M., Cassard, A. M., Wrzosek, L., Boschat, L., Bruneau, A., Ferrere, G., Puchois, V., Martin, J. C., Lepage, P., Le Roy, T., Lefèvre, L., Langelier, B., Cailleux, F., González-Castro, A. M., Rabot, S., Gaudin, F., Agostini, H., Prévot, S., Berrebi, D., … Perlemuter, G. (2016). Intestinal microbiota contributes to individual susceptibility to alcoholic liver disease. Gut, 65(5), 830–839. https://doi.org/10.1136/gutjnl-2015-310585

Lukić, I., Getselter, D., Koren, O., & Elliott, E. (2019). Role of Tryptophan in Microbiota-Induced Depressive-Like Behavior: Evidence From Tryptophan Depletion Study. Frontiers in Behavioral Neuroscience, 13, 123. https://doi.org/10.3389/fnbeh.2019.00123

McHugh, R. K., & Weiss, R. D. (2019). Alcohol Use Disorder and Depressive Disorders. Alcohol Research : Current Reviews, 40(1), arcr.v40.1.01. https://doi.org/10.35946/arcr.v40.1.01

Milstein, H. J., Bloomer, J. R., & Klatskin, G. (1976). Serum bile acids in alcoholic liver disease. Comparison with histological features of the disease. The American Journal of Digestive Diseases, 21(4), 281–285. https://doi.org/10.1007/BF01071839

Odlaug, B. L., Gual, A., DeCourcy, J., Perry, R., Pike, J., Heron, L., & Rehm, J. (2016). Alcohol Dependence, Co-occurring Conditions and Attributable Burden. Alcohol and Alcoholism, 51(2), 201–209. https://doi.org/10.1093/alcalc/agv088

Pessa-Morikawa, T., Husso, A., Kärkkäinen, O., Koistinen, V., Hanhineva, K., Iivanainen, A., & Niku, M. (2022). Maternal microbiota-derived metabolic profile in fetal murine intestine, brain and placenta. BMC Microbiology, 22(1), 46. https://doi.org/10.1186/s12866-022-02457-6

Phua, L. C., Wilder-Smith, C. H., Tan, Y. M., Gopalakrishnan, T., Wong, R. K., Li, X., Kan, M. E., Lu, J., Keshavarzian, A., & Chan, E. C. Y. (2015). Gastrointestinal Symptoms and Altered Intestinal Permeability Induced by Combat Training Are Associated with Distinct Metabotypic Changes. Journal of Proteome Research, 14(11), 4734–4742. https://doi.org/10.1021/acs.jproteome.5b00603

Rutledge, G. A., Sandhu, A. K., Miller, M. G., Edirisinghe, I., Burton-Freeman, B. B., & Shukitt-Hale, B. (2021). Blueberry phenolics are associated with cognitive enhancement in supplemented healthy older adults. Food & Function, 12(1), 107–118. https://doi.org/10.1039/d0fo02125c

Sullivan, L. E., Fiellin, D. A., & O’Connor, P. G. (2005). The prevalence and impact of alcohol problems in major depression: A systematic review. The American Journal of Medicine, 118(4), 330–341. https://doi.org/10.1016/j.amjmed.2005.01.007

Swendsen, J. D., Merikangas, K. R., Canino, G. J., Kessler, R. C., Rubio-Stipec, M., & Angst, J. (1998). The comorbidity of alcoholism with anxiety and depressive disorders in four geographic communities. Comprehensive Psychiatry, 39(4), 176–184. https://doi.org/10.1016/s0010-440x(98)90058-x

Trinchet, J. C., Gerhardt, M. F., Balkau, B., Munz, C., & Poupon, R. E. (1994). Serum bile acids and cholestasis in alcoholic hepatitis. Relationship with usual liver tests and histological features. Journal of Hepatology, 21(2), 235–240. https://doi.org/10.1016/s0168-8278(05)80401-5

van Sumere, C. F., Teuchy, H., Pé, H., Verbeke, R., & Bekaert, J. (1969). Quantitative investigation on the hippuric acid formation in healthy and diseased individuals. Clinica Chimica Acta, 26(1), 85–88. https://doi.org/10.1016/0009-8981(69)90289-7

Voutilainen, T., & Kärkkäinen, O. (2019). Changes in the Human Metabolome Associated With Alcohol Use: A Review. Alcohol and Alcoholism, 54(3), 225–234. https://doi.org/10.1093/alcalc/agz030

Vuong, H. E., Pronovost, G. N., Williams, D. W., Coley, E. J. L., Siegler, E. L., Qiu, A., Kazantsev, M., Wilson, C. J., Rendon, T., & Hsiao, E. Y. (2020). The maternal microbiome modulates fetal neurodevelopment in mice. Nature, 586(7828), 281–286. https://doi.org/10.1038/s41586-020-2745-3

Williams, H. R., Cox, I. J., Walker, D. G., Cobbold, J. F., Taylor-Robinson, S. D., Marshall, S. E., & Orchard, T. R. (2010). Differences in gut microbial metabolism are responsible for reduced hippurate synthesis in Crohn’s disease. BMC Gastroenterology, 10(1), 108. https://doi.org/10.1186/1471-230X-10-108

Winter, G., Hart, R. A., Charlesworth, R. P. G., & Sharpley, C. F. (2018). Gut microbiome and depression: What we know and what we need to know. Reviews in the Neurosciences, 29(6), 629–643. https://doi.org/10.1515/revneuro-2017-0072

Würtz, P., Cook, S., Wang, Q., Tiainen, M., Tynkkynen, T., Kangas, A. J., Soininen, P., Laitinen, J., Viikari, J., Kähönen, M., Lehtimäki, T., Perola, M., Blankenberg, S., Zeller, T., Männistö, S., Salomaa, V., Järvelin, M.-R., Raitakari, O. T., Ala-Korpela, M., & Leon, D. A. (2016). Metabolic profiling of alcohol consumption in 9778 young adults. International Journal of Epidemiology, 45(5), 1493–1506. https://doi.org/10.1093/ije/dyw175

Yilmaz, A., Ugur, Z., Bisgin, H., Akyol, S., Bahado-Singh, R., Wilson, G., Imam, K., Maddens, M. E., & Graham, S. F. (2020). Targeted Metabolic Profiling of Urine Highlights a Potential Biomarker Panel for the Diagnosis of Alzheimer’s Disease and Mild Cognitive Impairment: A Pilot Study. Metabolites, 10(9), E357. https://doi.org/10.3390/metabo10090357

Zheng, P., Chen, J., Huang, T., Wang, M., Wang, Y., Dong, M., Huang, Y., Zhou, L., & Xie, P. (2013). A Novel Urinary Metabolite Signature for Diagnosing Major Depressive Disorder. Journal of Proteome Research, 12(12), 5904–5911. https://doi.org/10.1021/pr400939q

Zheng, P., Wang, Y., Chen, L., Yang, D., Meng, H., Zhou, D., Zhong, J., Lei, Y., Melgiri, N. D., & Xie, P. (2013). Identification and validation of urinary metabolite biomarkers for major depressive disorder. Molecular & Cellular Proteomics: MCP, 12(1), 207–214. https://doi.org/10.1074/mcp.M112.021816

Zheng, P., Zeng, B., Zhou, C., Liu, M., Fang, Z., Xu, X., Zeng, L., Chen, J., Fan, S., Du, X., Zhang, X., Yang, D., Yang, Y., Meng, H., Li, W., Melgiri, N. D., Licinio, J., Wei, H., & Xie, P. (2016). Gut microbiome remodeling induces depressive-like behaviors through a pathway mediated by the host’s metabolism. Molecular Psychiatry, 21(6), 786–796. https://doi.org/10.1038/mp.2016.44

Zheng, Y., Beck, O., & Helander, A. (2011). Method development for routine liquid chromatography– mass spectrometry measurement of the alcohol biomarker phosphatidylethanol (PEth) in blood. Clinica Chimica Acta, 412(15–16), 1428–1435. https://doi.org/10.1016/j.cca.2011.04.022

