## Supplementary table 1 for "Alcohol use associated alterations in the circulating metabolite profile in the general population and in individuals with major depressive disorder"

**Supplementary table 1:** Correlations between self-reported alcohol intake and metabolite concentrations

| | General population cohort | | | | MDD cohort | | | |
|---|---|---|---|---|---|---|---|---|
| | Model 1 | | Model 2 | | Model 1 | | Model 2 | |
| Metabolite | β | p | β | p | β | p | β | p |
| 1-Methylhistamine | 0.01 | 0.8265 | | | 0.02 | 0.8394 | | |
| 2-Aminoisobutyric acid | 0.11 | 0.0937 | | | 0.20 | **0.0491** | 0.22 | **0.0336** |
| 2-Deoxycytidine | -0.03 | 0.6119 | | | 0.05 | 0.6280 | | |
| 3-Hydroxyanthranillic acid | -0.05 | 0.4247 | | | 0.09 | 0.3546 | | |
| 4-Pyridoxic acid | -0.06 | 0.3276 | | | 0.16 | 0.1232 | | |
| 5-Hydroxyindole-3-acetic acid | -0.17 | **0.0076** | -0.16 | **0.0183** | -0.04 | 0.6685 | | |
| Acetylcarnitine C2:0 | 0.07 | 0.3000 | | | 0.18 | 0.0709 | | |
| Adenosine | 0.00 | 0.9936 | | | -0.12 | 0.2481 | | |
| Alanine | -0.08 | 0.2054 | | | 0.10 | 0.3302 | | |
| Allantoin | 0.01 | 0.8586 | | | -0.07 | 0.4676 | | |
| Aminoadipic acid | -0.04 | 0.5508 | | | 0.04 | 0.7052 | | |
| Arginine | -0.08 | 0.2063 | | | -0.02 | 0.8428 | | |
| Asparagine | -0.05 | 0.4047 | | | -0.08 | 0.4248 | | |
| Aspartate | -0.01 | 0.9225 | | | -0.13 | 0.2099 | | |
| Asymmetric dimethylarginine | -0.08 | 0.2128 | | | 0.14 | 0.1693 | | |
| cAMP | -0.06 | 0.3465 | | | 0.04 | 0.7028 | | |
| Carnitine | 0.04 | 0.4778 | | | -0.03 | 0.7945 | | |
| Carnosine | -0.12 | 0.0641 | | | -0.21 | **0.0400** | -0.21 | **0.0348** |
| Chenodeoxycholic acid | 0.23 | **0.0003** | 0.19 | **0.0049** | 0.10 | 0.3399 | | |
| Cholic acid | -0.05 | 0.4693 | | | -0.14 | 0.1600 | | |
| Choline | -0.08 | 0.1792 | | | -0.01 | 0.9500 | | |
| Citrulline | 0.00 | 0.9968 | | | 0.03 | 0.7641 | | |
| Cotinine | 0.14 | **0.0253** | 0.03 | 0.5005 | 0.01 | 0.9323 | | |
| Creatine | -0.04 | 0.4963 | | | 0.05 | 0.6547 | | |
| Creatinine | -0.02 | 0.7372 | | | -0.08 | 0.4222 | | |
| Cystathionine | -0.07 | 0.2408 | | | 0.13 | 0.2080 | | |
| Cytidine | 0.18 | **0.0043** | 0.22 | **0.0011** | 0.21 | **0.0350** | 0.21 | **0.0388** |
| Cytosine | 0.05 | 0.3924 | | | -0.03 | 0.7976 | | |
| Decanoylcarnitine | 0.06 | 0.3142 | | | -0.10 | 0.3437 | | |
| D-Glucuronic acid | 0.03 | 0.5956 | | | 0.10 | 0.3441 | | |
| Dimethylglycine | -0.11 | 0.0854 | | | -0.06 | 0.5582 | | |
| FolicAcid | -0.06 | 0.3691 | | | 0.07 | 0.4919 | | |
| GABA | -0.01 | 0.8646 | | | 0.23 | **0.0205** | 0.25 | **0.0186** |
| gamma-Glutamylcysteine | -0.12 | 0.0662 | | | -0.13 | 0.2097 | | |
| Glutamine | -0.14 | **0.0312** | -0.20 | **0.0040** | -0.12 | 0.2398 | | |
| Glyceraldehyde | 0.02 | 0.7823 | | | 0.02 | 0.8670 | | |
| Glycine | -0.18 | **0.0044** | -0.12 | 0.0785 | -0.19 | 0.0543 | | |
| Glycine betaine | -0.05 | 0.4204 | | | 0.00 | 0.9904 | | |
| Glycocholic acid | 0.03 | 0.5855 | | | 0.07 | 0.5195 | | |
| Guanidinoacetic acid | 0.09 | 0.1742 | | | -0.10 | 0.3315 | | |
| Guanosine | -0.04 | 0.5105 | | | 0.10 | 0.3315 | | |
| Hexanoylcarnitine | 0.04 | 0.5547 | | | -0.04 | 0.6668 | | |
| Hippuric acid | -0.24 | **0.0001** | -0.19 | **0.0037** | -0.17 | 0.0882 | | |
| Histidine | -0.14 | **0.0244** | -0.20 | **0.0032** | -0.02 | 0.8088 | | |
| Homocysteine | -0.06 | 0.3380 | | | 0.11 | 0.2583 | | |
| Homogentisic acid | 0.12 | 0.0537 | | | -0.01 | 0.9178 | | |
| Homoserine | 0.02 | 0.7033 | | | 0.17 | 0.0963 | | |
| Hydroxyproline | 0.03 | 0.6795 | | | 0.15 | 0.1385 | | |
| Hypoxanthine | -0.04 | 0.5769 | | | 0.09 | 0.3953 | | |
| IMP | -0.01 | 0.8774 | | | -0.08 | 0.4170 | | |
| Inosine | -0.01 | 0.8377 | | | 0.11 | 0.3001 | | |
| Isobutyrylcarnitine C4:0 | -0.01 | 0.8942 | | | 0.03 | 0.7567 | | |
| Isoleucine | -0.04 | 0.5712 | | | 0.10 | 0.3412 | | |
| Isovalerylcarnitine C5:0 | 0.10 | 0.1274 | | | -0.08 | 0.4408 | | |
| Kynurenic acid | 0.02 | 0.7275 | | | -0.01 | 0.9213 | | |
| Leucine | 0.07 | 0.2561 | | | -0.01 | 0.9141 | | |
| L-Glutamic acid | 0.11 | 0.0789 | | | 0.14 | 0.1745 | | |
| L-Kynurenine | -0.07 | 0.2894 | | | 0.01 | 0.9513 | | |
| Lysine | -0.11 | 0.0958 | | | -0.02 | 0.8176 | | |
| Methionine | -0.08 | 0.2159 | | | -0.06 | 0.5641 | | |
| Niacinamide | 0.00 | 0.9749 | | | -0.06 | 0.5535 | | |
| Normetanephrine | 0.01 | 0.8877 | | | 0.00 | 0.9771 | | |
| Octanoylcarnitine | 0.02 | 0.7120 | | | -0.10 | 0.3232 | | |
| Ornithine | -0.04 | 0.4991 | | | 0.03 | 0.7744 | | |
| Pantothenicacid | 0.01 | 0.9069 | | | 0.03 | 0.7839 | | |
| Phenylalanine | -0.01 | 0.8790 | | | -0.07 | 0.4786 | | |
| Phosphoethanolamine | -0.06 | 0.3451 | | | -0.03 | 0.7874 | | |
| Proline | -0.08 | 0.1851 | | | 0.06 | 0.5338 | | |
| Propionylcarnitine | -0.04 | 0.5518 | | | -0.02 | 0.8156 | | |
| Ribose-5-phosphate | -0.12 | 0.0611 | | | -0.11 | 0.2579 | | |
| Serine | -0.14 | **0.0312** | -0.16 | **0.0218** | -0.15 | 0.1379 | | |
| Sorbitol | -0.05 | 0.3960 | | | 0.06 | 0.5693 | | |
| Spermidine | -0.06 | 0.3296 | | | 0.03 | 0.7664 | | |
| Succinate | -0.04 | 0.5094 | | | -0.03 | 0.7611 | | |
| Sucrose | -0.03 | 0.6277 | | | -0.09 | 0.3916 | | |
| Symmetric dimethylarginine | -0.06 | 0.3807 | | | 0.12 | 0.2215 | | |
| Taurine | -0.01 | 0.8572 | | | -0.10 | 0.3081 | | |
| Taurochenodeoxycholic acid | 0.06 | 0.3314 | | | 0.13 | 0.1917 | | |
| Taurocholic acid | 0.15 | **0.0148** | 0.22 | **0.0012** | 0.14 | 0.1537 | | |
| Threonine | 0.02 | 0.7947 | | | 0.04 | 0.7082 | | |
| Trimethylamine-N-oxide (TMAO) | -0.03 | 0.6164 | | | -0.07 | 0.4718 | | |
| Tryptophan | -0.07 | 0.2948 | | | 0.00 | 0.9719 | | |
| Tyrosine | 0.01 | 0.8567 | | | 0.18 | 0.0728 | | |
| Valine | -0.01 | 0.8626 | | | -0.01 | 0.9461 | | |
| Xanthine | 0.07 | 0.2993 | | | 0.15 | 0.1470 | | |
| Xanthosine | -0.03 | 0.6718 | | | -0.34 | **0.0006** | -0.38 | **0.0002** |

Legend: β, standardized β coefficient from linear regression model; Model 1, linear regression without confounders; Model 2, linear regression model with confounders (age, sex, use of vegetables, use of fruits, smoking, BMI, education level, and cohabitation status)
